# Development and validation of an all-in-one Bat-Clade genomic sequencing and host identification protocol

**DOI:** 10.1101/2024.10.31.24316523

**Authors:** Fernanda Marques de Souza Godinho, Aline Campos, Rosana Huff, Amanda Pellenz Ruivo, Thales Bermann, Milena Bauerman, Franciellen Machado dos Santos, Taina Machado Selayaran, Artur Beineke Correa, Raissa Nunes dos Santos, Paulo Michel Roehe, Gabriel da Luz Wallau, Richard Steiner Salvato

## Abstract

Rabies virus (RABV), a fatal zoonotic pathogen, remains a significant public health concern, with bat-maintained lineages accounting for all currently documented cases in Brazil. Despite the availability of pharmacological prophylaxis for humans and animals, the high genetic diversity of RABV in diverse natural bat hosts and continued circulation in multiple animals pose challenges for effective surveillance. Here, we developed and validated a novel, rapidly deployable amplicon-based sequencing approach for rabies virus (RABV) genomic surveillance. This “all-in-one” protocol integrates whole RABV genome sequencing with host species identification through COI gene amplification and sequencing, addressing the challenges posed by RABV’s high genetic diversity and complex transmission dynamics. We assessed the protocol’s effectiveness by sequencing 25 near-complete RABV genomes from host species across four distinct families (Bovidae, Equidae, Felidae, and Microchiroptera) obtained from the Rabies Control and Surveillance Program in Rio Grande do Sul State, Southern Brazil. The method achieved an average genome coverage of 91.4% at a minimum 5x read depth, with a mean depth coverage of 816x across sequenced genomes. The results demonstrated significant Bat-Clade sublineage diversity, which was classified using the MADDOG RABV lineage system. The protocol successfully identified three bat species (*Tadarida brasiliensis, Desmodus rotundus*, and *Myotis nigricans*) among the samples, highlighting its capability for precise host identification. This study presents a powerful tool for high-resolution evaluation of RABV genomic features and host identification, enabling more targeted public health interventions. This new approach has the potential to enhance RABV surveillance capabilities, contributing to more effective rabies control strategies within a One Health framework.

## Introduction

Rabies virus (RABV), a neurotropic *Lyssavirus* of the *Rhabdoviridae* family, is a zoonotic pathogen capable of infecting several mammalian species, including humans, domestic animals (e.g., dogs, cats, livestock), and diverse wildlife (Fooks *et al.*, 2014; Rupprecht *et al.*, 2018). Despite a World Health Organization (WHO) initiative to end human deaths from dog-mediated rabies by 2030 (WHO, 2018), this disease remains a significant public health concern, claiming an estimated 59,000 lives annually and resulting in billions of dollars in economic losses globally (Hampson *et al.* 2015). In the Americas, where dog-mediated rabies is mainly controlled, hematophagous bats have emerged as the primary source of human rabies (Velasco-Villa *et al.*, 2017). RABV isolates are classified into two major phylogenetic groups: bat-related and dog-related, with the bat-related group primarily found in the Americas (Badrane & Tordo, 2001). Neotropical bats are considered the main wild animal reservoirs of RABV, supported by the higher genetic diversity and ancestral state of bat-clade lineages (Mollentze *et al.,* 2014; Oliveira *et al.*, 2020; Velasco-Villa *et al.*, 2017).

Recent genomic studies have revealed complex evolutionary dynamics within the bat-related RABV group. Troupin *et al*. (2016) found no correlation between root-to-tip geneticdistance and sampling time in the bat-related RABV group, nor when combined with the dog-related RABV group, indicating that these viruses have not evolved in a clock-like manner. This observation, combined with the substantial rate variation already observed in bat-associated RABV, underscores the need for advanced genomic surveillance techniques to understand viral evolution and transmission dynamics better (da Cunha *et al.*, 2023). In Brazil, despite significant reductions in urban human rabies cases, the sylvatic cycle driven by bat-clade RABV remains a primary source of animal and human infections, highlighting the ongoing challenges in controlling rabies (Campos *et al.*, 2020). According to the Ministry of Health, from 2019 to 2023 in Brazil, 11 cases of human rabies and 78 cases of animal rabies were recorded, 8 and 29 bat-associated, respectively (MS, 2024).

Climate and environmental changes are rapidly altering wildlife communities, creating the potential for new species interactions and pathogen spillover. This urgency underscores the news for immediate action to prevent the emergence of new reservoir species and shifts in pathogen transmission dynamics. In the future, different wildlife species could become RABV reservoirs or develop the ability to transmit the virus to other animals, including humans (Escobar *et al.*, 2023; Mollentze *et al.*, 2014). Genomic surveillance of zoonotic pathogens, including RABV, holds immense potential to provide high-resolution insights into viral evolution, mutation tracking, spillover events, and wildlife reservoir maintenance, which are crucial for informing effective mitigation strategies (Campbell *et al.*, 2022; Escobar *et al.*, 2023; Gigante *et al*., 2020; Kuzmin *et al.*, 2012). RABV surveillance in Brazil is mainly performed by classical, labor-intensive methods like RT-PCR and targeted sequencing of viral genes. These approaches have limitations in detecting genome-wide mutations and definitively identifying host species (Cappelari *et al*., 2022; Oliveira *et al.*, 2010).

Recent advancements in whole genome sequencing (WGS) offer more data for high resolution phylogenetic analysis and can differentiate between samples that appear identical at the partial genome level (Campbell *et al*., 2022; Hyeon *et al*., 2021). While these methods effectively detect and type RABV, they often fail to identify the host species definitively, particularly when the sample quality is compromised or the host species is not visually identifiable. Integrating cytochrome oxidase I (COI) gene sequencing into surveillance protocols has been proposed to address this limitation. This approach facilitates precise host identification and enhances the overall effectiveness of rabies surveillance (Hebert *et al.*, 2003). Given the limited understanding of this high-impact zoonotic virus from a genomic perspective, it is essential to establish cost-effective RABV genomic surveillance protocols to substantiate public health surveillance and mitigation programs (Campbell *et al.*, 2022). Additionally, despite the importance of the RABV zoonotic cycle, complete genome sequence data remain limited for RABV lineages circulating in bats, particularly in regions like Brazil (Oliveira *et al*., 2020).

To address these gaps, we developed a novel, rapidly deployable, and flexible amplicon-based sequencing approach usable with protocols widely established during the COVID-19 pandemic and suitable for different host identification that can recover near complete RABV genomes and precisely identify the original host. This “all-in-one” protocol can generate actionable surveillance results within a One Health framework. This interconnected approach is crucial for monitoring and controlling zoonotic pathogens like RABV (Campbell *et al*., 2022; Wadhwa *et al.*, 2017).

## Methods

### Samples

Samples were collected through the passive Rabies Surveillance Program in Rio Grande do Sul, Brazil, a collaboration between the State Agriculture, Livestock, and Rural Department (SEAPDR) and the State Health Department. Central Nervous System (CNS) samples were sourced from three primary sample types: deceased domestic herbivores showing neurological signs, deceased companion animals with neurological signs or those that died during post-exposure observation, and any wild mammals found dead. These samples were part of routine diagnostics for potential rabies cases, exempt from Ethics Commission on Animal Use (CEUA) approval.

### RABV molecular detection

A total of 200 uL of homogenate sample, containing 50 mg of brain tissue fragments macerated in Basal Medium Eagle (BME), were added by 800μl of TRIzol™ Reagent (Invitrogen, Grand Island, NY, USA) in 2.0 ml microtubes. The tissue was disrupted using a vortex at maximum speed for 15 seconds, followed by room incubation for 5 min and 180 µl of chloroform addition. The mixture was mixed for 15 seconds, incubated at room temperature for 3 min, and centrifuged at 12,000 RFC at 4°C for 15 minutes. Viral RNA extraction was carried out with 200 μl of supernatant using a commercially available Extracta Kit Fast – DNA e RNA Viral (MVXA-PV96-B FAST) and the Loccus Extracta® 96 equipment (Loccus, Sao Paulo, Brazil) following the manufacturer’s instructions. Extracted RNA samples were stored at ™80°C until tested by RT-qPCR.

The TaqMan assay single-step RT-PCR assay was conducted on BioRad CFX96 PCR System (California, CA, USA) using Invitrogen™ SuperScript™ III One-Step RT-PCR System (Thermo Fisher Scientific, Waltham, MA, USA). The modified version of pan-Lyssavirus TaqMan RT-qPCR reaction mix consisted of 12.5 μl of 2x Reaction Mix, 1 μl of SuperScript™ III RT/Platinum™ Taq Mix, 2 μl of extracted RNA, 400 nM of forward and reverse primers for the LN34, 100 nM of probe and nuclease-free water to a final volume of 25 μl. For the amplification of the β-actin gene, the concentrations of primers and probes were 100 nM; the remaining reagents were the same. For both targets, the amplification protocol was 30 min at 50°C, then 10 min at 95°C, followed by 45 cycles of denaturation at 95°C for 15 s and annealing/extension at 56°C for 45 s (Wadhwa *et al.*, 2017). Reactions targeting LN34 were tested in triplicates in separate wells from the β-actin target (a list of primers and probes used in this study are available in the supplementary material).

### Primers design

To find the closer RABV genomes from those circulating in Southern Brazil, we reconstructed a phylogenetic tree with seventy-four RABV partial nucleoprotein (N) gene sequences from Southern Brazil, along with a global dataset with 320 sequences covering all RABV clades. An amplicon of 815 nucleotides from the N gene was obtained using sense primer 21G and antisense primer P784, as described by Capellari *et al*. (2022). The fragments were sequenced by the Sanger method, using the BigDye Terminator v3.1 Cycle Sequencing Kit (ThermoFisher) and a 3500xl Genetic Analyzer (Applied Biosystems).

The 320 WGS sequences described by Troupin *et al.* (2016) were downloaded from NCBI and aligned using MAFFT with the FFT-NS-2 algorithm, and then we used the MAFFT --addfragments option to align N gene fragments to WGS sequences alignment. Next, we used IQTree2 to find the best nucleotide substitution model and reconstruct a phylogenetic tree from the alignment file. Then, we retrieve the complete genome sequences that clustered with the partial N sequences in the same clades. The following sequences (AB519641, KX148100, KX148269, JQ685947) were aligned and used as input to primer design using PrimalScheme (https://primalscheme.com), defining an amplicon size range from 760 to 840bp.

The resulting multiplex primer scheme was aligned to WGS sequences used in PrimalScheme and our N gene sequences. Then, we manually degenerated the primer set to increase the chances of capturing the RABV diversity from different bat subclades. Finally, we ended up with 23 forward primers and 24 reverse primers distributed into two primer pools. We also amplified a fragment of the mitochondrial gene *cytochrome c oxidase subunit I* (COI), designed for general vertebrates for bat species identification. The primers used vertCOI_7194 (5’-CGM ATR AAY AAY ATR AGC TTC TGA Y -3’) and Mod_repCOI_R (5’-TTC DGG RTG NCC RAA RAA TCA -3’) were described by Reeves *et al.* (2018), and generates a fragment of 395 base pairs. The primers for gene COI amplification were also added to the primer pool previously cited, enabling both RABV sequencing and host identification in a single assay.

### Next-generation sequencing

To develop an easy-to-use and rapidly deployable protocol, we adapted the Illumina COVISeq Assay by replacing the SARS-CoV-2 primers pools for RABV+COI primers, designed as described above. We followed the Illumina COVIDSeq RUO Kits Reference Guide. First, we annealed 8.5 µl from extracted RNA with 8.5 µl Elution Prime Fragment 3HC Mix and incubated at 65°C for 3 minutes. To synthesize the first strand cDNA, we prepared a master mix with First Strand Mix (9 µl) and Reverse Transcriptase (1 µl), and then 8 µl of the master mix was added to previously annealed RNA and incubated at 25°C for 5 minutes, 50°C for 10 minutes and 80°C for 5 minutes. The PCR reactions to amplify cDNA were performed using two mixes with 15 µl Illumina PCR Mix, 4.3 µl of each primer pool, and 4.7 µl of Nuclease-free water. Thermal cycler conditions were 98°C for 3 minutes, allowed by 35 cycles of 98°C for 15 seconds and 63°C for 5 minutes. The protocol was also validated using other reagent kits (Q5® High-Fidelity DNA Polymerase and Invitrogen Platinum™ Taq DNA Polymerase). See complete protocols in supplementary files.

Agarose gel electrophoresis was used to check the amplicons before proceeding to library constructions. Amplicons visualized at agarose gel were used for library preparation according to Illumina COVIDSeq for Illumina DNA Prep RUO Kits Reference Guide and then sequenced at platform Illumina MiSeq using Reagent Kit v3 (600-cycle). The detailed step-by-step protocol is fully available at https://www.protocols.io/view/rabies-virus-bat-clade-sequencing-8epv5x3bng1b/v2.

### Genome assembly

Fastq raw reads files were used as input into ViralFlow 1.0 Pipeline (https://viralflow.github.io) (Dezordi *et al.*, 2022; da Silva *et al.*, 2024) for reference-based assembly, variant calling, and consensus generation using the following reference genomes: AB519641 for Bats DR clade, JQ685947 for Bats LC clade and EU293116 for Bats TB2. Improved coverage breadth and depth were obtained when using the most similar subclade-specific references instead of a single bat-clade reference genome due to the high genetic diversity of RABV from these clades. So, we first assembled the sequences using the bat clade reference (JQ685956). We ran the consensus resulting sequence at the RABV-GLUE typing tool (http://rabv-glue.cvr.gla.ac.uk/), and then we took the nearest reference from the database and reassembled using this closest relative reference genome cited above. The ViralFlow pipeline provided all mapping, coverage, and depth quality values along with consensus sequencing fasta files that were retrieved for posterior clade assignment and phylogenetic analysis.

The COI fragment was assembled using a de novo assembly approach with SPAdes v3.15.4 (Prjibelski *et al.*, 2020). Contigs were selected based on scaffolds ranging from 385 to 405 bp, focusing on recovering the 395 bp COI fragment enriched in the initial primer pool for host identification. Once the COI fragments for each sample were assembled, they were queried against the BOLD (Barcode of Life Data) System database (Ratnasingham & Hebert, 2007) for taxonomic classification.

### RABV lineage assignment and phylogenetic analysis

RABV clades were designated by the RABV-GLUE website (http://rabv.glue.cvr.ac.uk) using the file with consensus fasta sequences. In addition, we implemented the MADDOG pipeline (Campbell *et al.*, 2022) (https://github.com/KathrynCampbell/RABV_Lineages) for lineage assignment. We retrieved the RABV Bat-clade sequences from Troupin *et al.* (2016) for phylogenetic analysis. The dataset within our samples was aligned using MAFFT (Katoh *et al.*, 2019). Subsequently, we employed IQ-TREE2 (Minh *et al*., 2020) to estimate a Maximum Likelihood (ML) phylogenetic tree. The analysis was conducted under the General Time Reversible (GTR) nucleotide substitution model (+F+I+G4), identified as the best-fit model by the ModelFinder application within IQ-TREE2 (Kalyaanamoorthy *et al*., 2017) using bootstrap along with SH-aLRT for assessing branch supports. The tree was then rooted using the Australian bat lyssavirus genome (NC_003243.1), the most recent ancestral sequence, and annotated using FigTree (Rambaut, 2018). We also generated an analysis of genomic divergence among the sequenced genomes using an in-house R script, that provides the pairwise sequence divergence estimation. The method calculates pairwise sequence divergence by determining the percentage of differing positions between two sequences while excluding gaps. This is expressed using the formula: divergence percentage equals the number of differences divided by the total number of valid positions, multiplied by 100.

## Data Availability

The data generated during this study, supplementary material, and all scripts implemented are available at https://github.com/salvatolab/RABV-Bat-clade-sequencing-protocol. RABV sequences generated are available on NCBI Virus under accession numbers *waiting accession number*.

## Results

### Initial validation

For the initial validation of the primer scheme designed, we sequenced the 25 RABV genomes from the same samples utilized to obtain the N partial sequence, following the proposed protocol. These samples were collected from different host species (Bovidae, Equidae, Felidae, Chiroptera) and, according to previous N gene clade assignment performed on RABV-GLUE, belonged to three different minor clades: TB2, DR, and one sequence was not classified to a minor clade.

The included samples showed Ct values ranging from 17 to 35 (average: 25.4), and we generated from 20,000 to 224,000 reads per sample (average: 116,000 reads), with an average mapped reads proportion of 98% (Figure 1). The average genome coverage was 91.4% (ranging from 54 to 100), considering a 5x minimum read depth coverage per position. The mean depth coverage among the sequenced genomes ranged from 22x to 1591x (average: 816x). A review of the genomic coverage identified two consistent gaps in the regions from 4,526 nt to 4,717 nt and 4,844 nt to 5,270 nt (in the genes coding for the RABV glycoprotein and the RNA-dependent RNA polymerase L protein, respectively), which were present in all sequenced lineages (Supplementary Figure 1). The host identification of the 25 samples tested had the following distribution: 16 bovids, two equids, one cat, and six bats. Mitochondrial gene COI allowed the classification of bats into three different species: *Tadarida brasiliensis* (3), *Desmodus rotundus* (2), and *Myotis nigricans* (1).

**Figure 1.**
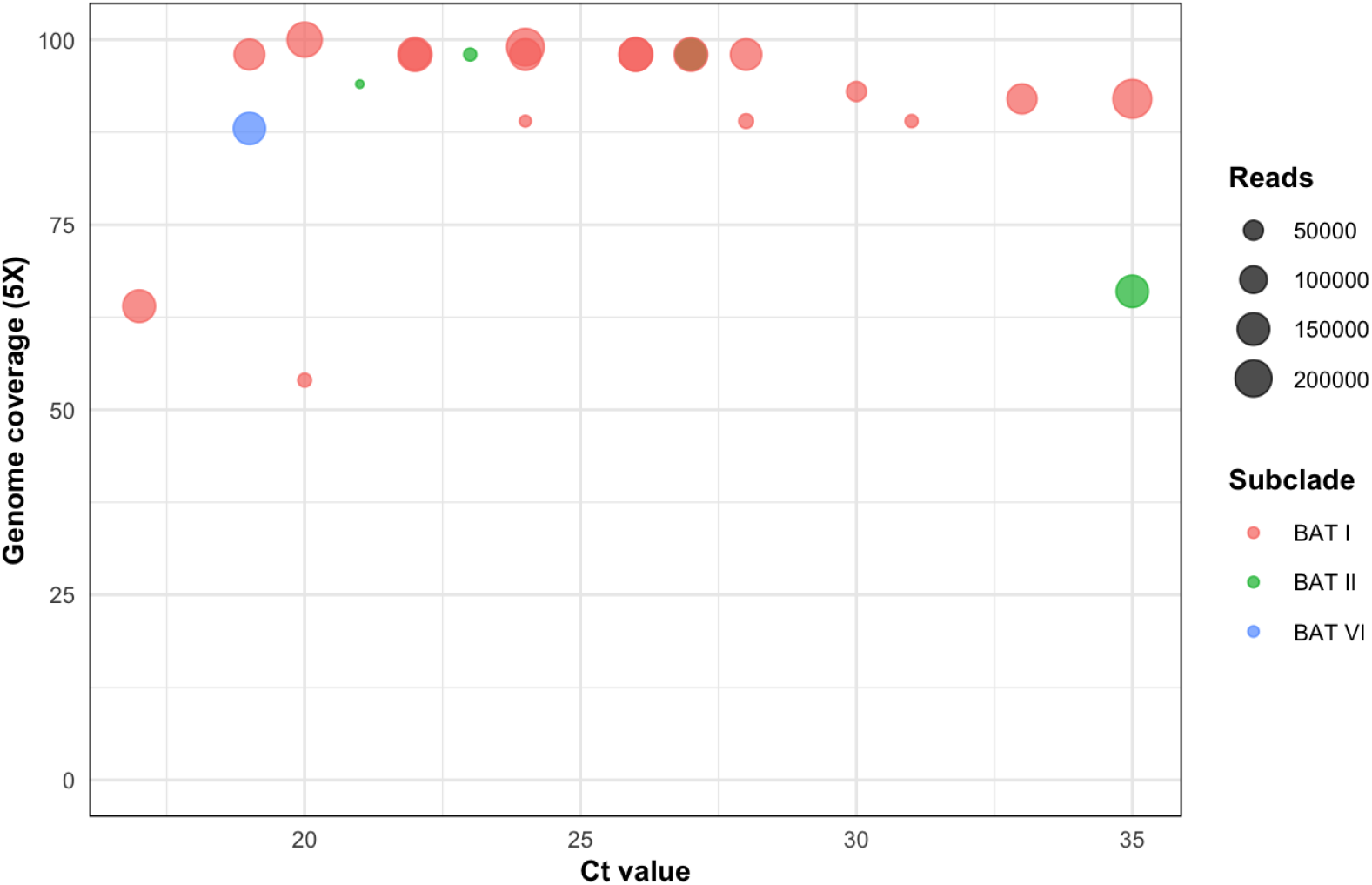
Coverage and depth metrics for 25 RABV genomes sequenced, along with the number of reads generated for each sample.

### Lineage assignment and phylogenetic analysis

According to RABV-GLUE lineage classification, from our 25 included genomes 20 (80%) were from the Bats DR clade, 4 (16%) from Bats TB2, and one genome RABV-GLUE was not able to assign a clade (even though the genome shows 80% coverage). MADDOG assignment shows a diversity of RABV sublineages: Bats DR_A1.1.2 (11/25), Bats DR_A1.1.7 (5/25), Bat_B1 (4/25), Bats DR_A1.1.4 (2/25), Bat_B1.1 (1/25), Bats_A1 (1/25) and Bats DR_M1 (1/25) (Table 1).

**Table 1.** Lineage Classification of RABV Genomes Based on RABV-GLUE and MADDOG Assignments.

| RABV-GLUE minor clade | MAD DOG lineage | Sequences |
| --- | --- | --- |
| Bats DR | Bats DR_A1.1.2 | 9 |
| Bats DR | Bats DR_A1.1.7 | 5 |
| Bats TB2 | Bat_B1 | 4 |
| Bats DR | Bats DR_A1.1.4 | 2 |
| Bats DR | Bat_B1.1 | 1 |
| Bats DR | Bats DR_M1 | 1 |
| Unknown | Bats_A1 | 1 |

We reconstructed a phylogenetic tree, including the 25 sequenced genomes by the Sanger method and our developed protocol, alongside 61 complete/near-complete RABV Bat-clade genomes worldwide (Troupin *et al*., 2016). This analysis aimed to provide a phylogenetic context for the RABV strains circulating in Rio Grande do Sul and the successful recovery of highly divergent viruses from different clades due to the degeneration of primer bases implemented at the primer scheme development stage. The sequences from this study were grouped into three (BAT I, II and VI) out of six monophyletic clades defined in this study (new clade definitions were added once RABV-GLUE reported non-monophyletic groups) (Figure 2). Clade BAT I comprised 20 sequences classified within the DR minor clade by RABV-GLUE. Clade BAT II included four sequences assigned within the TB minor clade. Clade BAT VI contained a unique sequence that could not be classified into any minor clade by RABV-GLUE. Notably, this sequence, unclassified by RABV-GLUE, was assigned to the Bats_A1 sublineage by MADDOG and was positioned as an outgroup to the bat clade in our phylogenetic tree.

**Figure 2.**
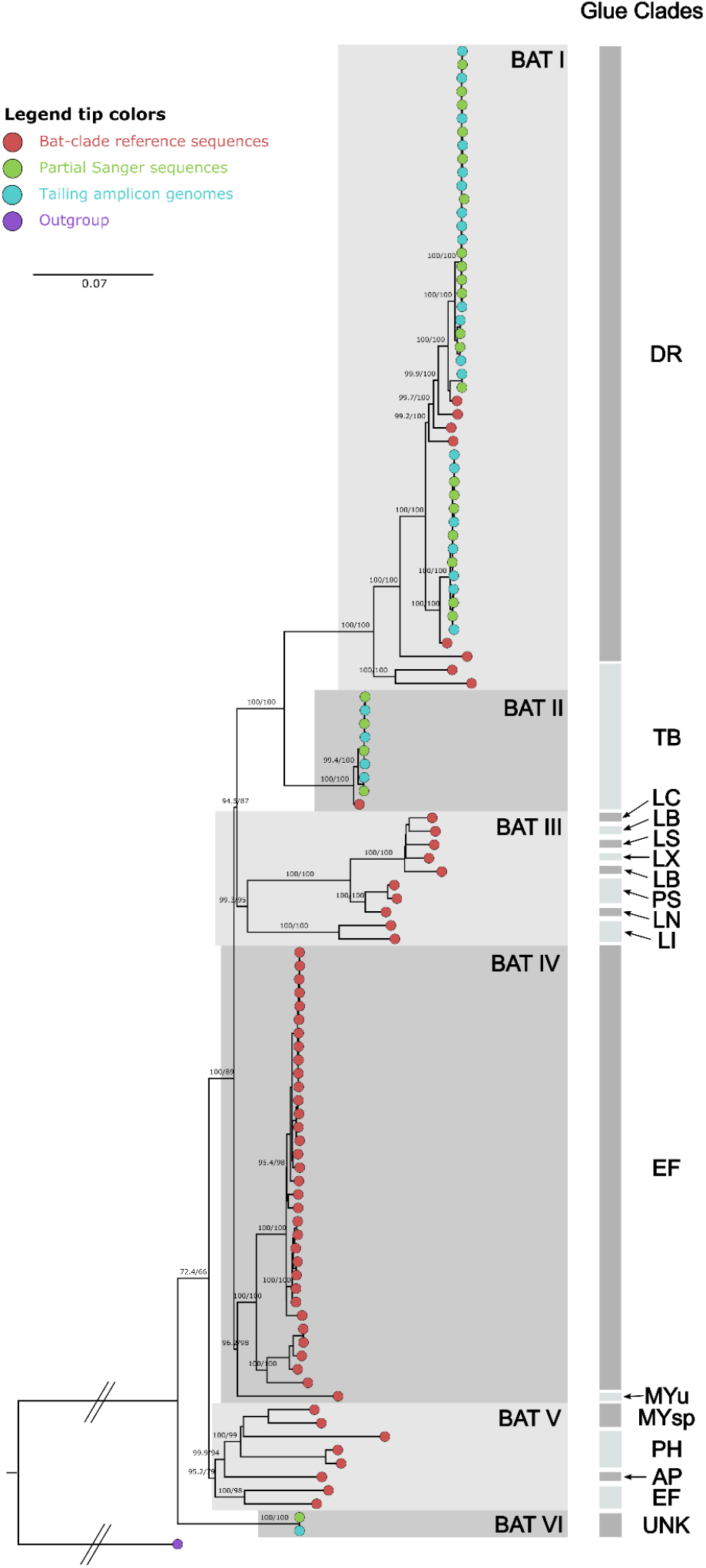
Maximum Likelihood phylogenetic tree including the sequences generated using the proposed protocol along the same samples Sanger sequences and Bat-clade reference sequences obtained from Troupin *et al.* 2016. The tip colors indicate the sequence source according to the legend. RABV-GLUE minor clade assignment is shown on the right bars. The values in the tree represent bootstrap support values.

In order to investigate the within and between clade divergence of RABVs from the BAT clades characterized, we performed a Hamming dissimilarity distance analysis (Figure 3). The intra-clade divergence ranged from 0-4% in BAT I and 0-1 in BAT II. BAT VI single genomic sequence diverged from 12-14% from all other clades. For inter-clade divergence, BAT I vs BAT II ranged from 10-11%, while BAT I vs BAT VI divergence was 14% and BAT II vs BAT VI was 12%. These results corroborate the large variability of bat clades circulating in their natural reservoir bat taxa in South America.

**Figure 3.**
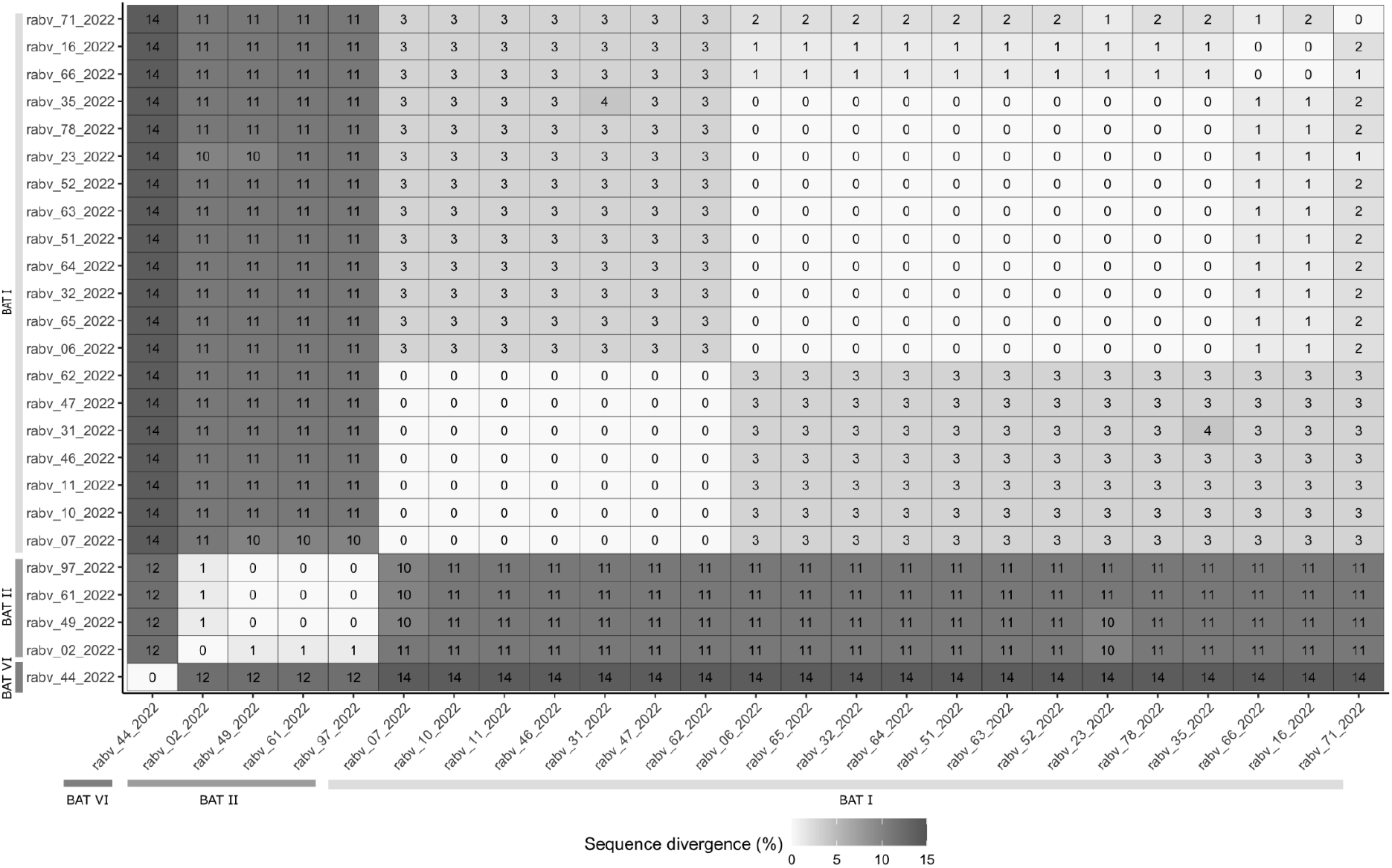
Heatmap illustrating the Hamming dissimilarity matrix (in percentage) among sequences generated by the tiling amplicon protocol. This heatmap visually represents the relative genetic differences, considering the rate of divergent nucleotide positions between each pair of sequences within our dataset.

## Discussion

Rabies virus (RABV) remains a highly lethal zoonotic pathogen with significant public health implications, particularly in the Americas, where bat-maintained lineages are predominant and continue to circulate and induce mortality among bats and wild/domestic animals (Fooks *et al.*, 2014; Velasco-Villa *et al*., 2017). Despite ongoing global efforts to control rabies, including the World Health Organization’s initiative to eliminate human deaths from dog-mediated rabies by 2030 (WHO, 2018), the disease poses substantial health and economic burdens (Hampson *et al.*, 2015). Our study presents a novel, rapidly deployable amplicon-based sequencing approach for RABV surveillance, integrating viral genomic characterization and host identification within a single assay. This comprehensive method addresses critical gaps in current RABV surveillance strategies and offers new insights into the diverse variants of RABV prevalent in southern Brazil and the broader southern region of the Americas.

The effectiveness of our protocol is demonstrated by its performance across a range of viral loads, as illustrated in Figure 1. Notably, we achieved successful genome sequencing across a broad spectrum of Ct values (17-35), encompassing samples with high and low viral loads. This robustness is crucial in field situations where sample quality can vary significantly, a challenge often faced in rabies surveillance (Cappelari *et al*., 2022). Our method achieved an average genome coverage of 91.4% at a minimum 5x read depth, with a mean depth coverage of 816x across sequenced genomes. This high-quality data enables confident identification of genetic variations and high resolution phylogenetic analysis, which is crucial for tracking RABV evolution and spread (Campbell *et al.*, 2022; Gigante *et al.*, 2020).

The high level of genetic diversity naturally infecting bats and corroborated by the data of this study underscores the importance of using whole genome sequencing approaches for RABV surveillance, as methods targeting only specific genes may miss significant variations across the genome (Kuzmin *et al*., 2012). Identifying consistent gaps in genomic coverage (4,526-4,717 nt and 4,844-5,270 nt) across all sequenced lineages warrants further investigation and may represent areas of high genetic variability or structural complexity that challenge current sequencing methods.

The integration of host identification through COI gene sequencing, as suggested by Hebert *et al.* (2003), proved valuable to our protocol. This approach addresses a critical challenge faced by rabies control teams in accurately identifying host species, particularly bats and other wild mammals, that may be difficult to distinguish morphologically (Mollentze *et al.*, 2014). The identification of three different bat species (*Tadarida brasiliensis, Desmodus rotundus*, and *Myotis nigricans*) among our samples highlights the diversity of RABV reservoirs and the importance of species-specific surveillance strategies, aligning with previous findings on the complexity of bat-associated RABV ecology (Badrane & Tordo, 2001; Troupin *et al.*, 2016).

Our amplicon-based sequencing approach is rapid and cost-effective and facilitates real-time monitoring, enabling proactive public health measures to prevent and control disease outbreaks. By adopting the widely-used Illumina COVISeq Assay, our protocol leverages existing infrastructure and expertise developed during the COVID-19 pandemic, making it more accessible to laboratories with limited resources (Quick *et al*., 2017). The successful integration of our protocol into the state rabies surveillance program of Rio Grande do Sul demonstrates its practical applicability in real-world settings, addressing the need for improved surveillance methods highlighted by previous studies (Oliveira *et al*., 2010).

Our study has some limitations, the consistent gaps in genomic coverage warrant further investigation and potential refinement of the primer design. Future research should focus on expanding the application of this method to larger sample sets and diverse geographic regions, potentially uncovering new insights into RABV evolution and transmission dynamics. The developed application of the technique is critical in the context of climate and environmental changes altering wildlife communities, as discussed by Mollentze *et al*. (2014) and Escobar *et al*. (2023).

Recent studies have highlighted the impact of climate change on bat populations and their associated pathogens (Escobar *et al.*, 2023). This environmental shift may alter RABV transmission dynamics, emphasizing the need for adaptive surveillance strategies. Furthermore, advancements in genomic sequencing technologies have enabled more precise phylogenetic analyses of RABV (Hyeon *et al.*, 2021; Oliveira *et al*., 2020), allowing for a better understanding of viral evolution and spread. These developments underscore the importance of our approach in providing a flexible and comprehensive tool for RABV surveillance that can adapt to changing ecological conditions and leverage emerging technologies.

In conclusion, our “all-in-one” protocol for RABV genomic surveillance represents a significant advancement in the field, offering a cost-effective and efficient method for simultaneous viral characterization and host identification. This approach aligns well with the One Health framework (Wadhwa *et al.*, 2017), providing actionable results that can inform targeted control and prevention strategies. By expanding genomic surveillance, the generated data will fill gaps in understanding the dispersion of targeted RABV lineages and precisely monitor potential spillover events, addressing concerns raised in previous studies (Zhang *et al.*, 2018). Our method addresses the limitations of classical, labor-intensive methods like RT-PCR and targeted sequencing of viral genes, as noted by Cappelari *et al*. (2022) and Oliveira *et al*. (2010), by offering a more comprehensive genomic perspective. This protocol can be crucial in informing public health strategies and contributing to global efforts to control and eliminate human rabies, supporting initiatives such as the WHO’s goal to end human deaths from dog-mediated rabies by 2030 (WHO, 2018).

## Data Availability

The data generated during this study, supplementary material, and all scripts implemented are available at https://github.com/salvatolab/RABV-Bat-clade-sequencing-protocol.

